# Are We Seeing the Unseen of Human Trafficking? A Retrospective Secondary Analysis of the CTDC K-Anonymized Global Victim of Trafficking Data Pool in the Period 2010-2020

**DOI:** 10.1101/2023.01.06.23284257

**Authors:** Ala’a B. Al-Tammemi, Asma Nadeem, Laila Kutkut, Manal Ali, Khadijah Angawi, Maram H. Abdallah, Rana Abutaima, Rasha Shoumar, Rana Albakri, Malik Sallam

## Abstract

**Background:** Human trafficking is considered a hidden global threat with unsubstantiated numbers despite the estimation of the presence of around 40.3 million victims worldwide. Human trafficking results in severe detrimental impacts on both mental and physical health. Given the sensitivity and negative consequences of human trafficking on the global system and victims, and considering the scarce research in this area, our current study aimed at describing the (i) Sociodemographic profiles of anonymized victims, (ii) Means of control, and (iii) Purpose of trafficking, utilizing the largest anonymized and publicly available dataset on victims of human trafficking.

**Methods:** This is a retrospective secondary analysis of the Counter-Trafficking Data Collaborative (CTDC) data pool in the period from 2010 to 2020. The utilized dataset is called the k-anonymized global victim of trafficking dataset, and it is considered the largest global dataset on victims of human trafficking. Data from the k-anonymized data pool were extracted and exported to Statistical Package for Social Sciences, SPSS® version 27.0 for Windows (IBM Corp. Version 27.0.Armonk, NY) for quality check and analysis using descriptive statistics.

**Results:** A total of 87003 victims of human trafficking were identified in the period from 2010 to 2020. The most age category encountered among victims was 9-17 years with around 10326 victims (11.9%), followed by 30-38 years with 8562 victims (9.8%). Females comprised 70% of the sample with 60938 victims. The United States (n=51611), Russia (n=4570), and the Philippines (n=1988) comprised the most countries of exploitation/trafficking. Additionally, the year 2019 witnessed the greatest number of victims registered for assistance by anti-trafficking agencies with around 21312 victims (24.5%). Concerning means of control, threats, psychological abuse, restriction of the victim’s movement, taking the victim’s earnings, and physical abuse were the most reported means. Around 42685 victims (49.1%) reported sexual exploitation as the purpose of their trafficking, followed by forced labor with 18176 victims (20.9%).

**Conclusion:** Various means and methods can be used by traffickers to control the victims to be trafficked for many purposes, with sexual exploitation and forced labor being the most common ones. Global anti-trafficking efforts should be brought together in solidarity through utilizing the paradigm of protection of victims, prosecution of traffickers, prevention of trafficking, and inter-sectoral partnerships. Despite being a global concern with various reports that tried to capture the number of trafficked victims worldwide, human trafficking still has many unseen aspects that impose a significant challenge and adds to the global burden in combatting this threat.

## 1. BACKGROUND

Generally, migration is defined as changing the place of habitual residence for a variety of reasons, and this can be temporarily or permanently, within the same country or through moving to another country (1,2). For simplification, migration can be classified into internal migration (within the same country), or external migration (moving to another country). Also, various geopolitical, technological, and environmental factors are shaping global migration (2). People migrate for a variety of reasons, some of which are man-made hazards (e.g., wars and armed conflicts, violation of human rights), natural/environmental disasters (earthquakes, floods, Tornados, epidemics, droughts), or other peaceful reasons such as studying, or looking for job opportunities (i.e., labor migration) (3). Statistically, around 3.6% of the world’s population are migrants in 2020 (i.e., people who migrated) and this corresponds to 281 million according to the most recent world migration report 2022 released by the International Organization for Migration (IOM) (2). Of the global 281 million migrants as of 2020, 135 million are female, and 146 are males, with around 169 million people migrating due to labor reasons (i.e., Labour migrants).

From a legal perspective, migration can be categorized into regular migration which is considered an authorized movement managed by various official authorities around the world, or irregular migration which corresponds to unauthorized movement through illegal ways such as smuggling and human trafficking (4). The term human trafficking is also known in the literature as trafficking in person (TIP) or modern-day slavery (5). The United Nations (UN) defines human trafficking as “*the recruitment, transportation, transfer, harbouring or receipt of persons, by means of the threat or use of force or other forms of coercion, of abduction, of fraud, of deception, of the abuse of power or of a position of vulnerability or of the giving or receiving of payments or benefits to achieve the consent of a person having control over another person, for the purpose of exploitation*” (6,7). Also, the US National Human Trafficking Resource Centre (NHTRC) defines human trafficking as “*Human trafficking involves the use of force, fraud, or coercion to compel a person into commercial sex acts or labour or services against his or her will*” (8). Human trafficking is considered a crime and a severe multidimensional violation of human rights as well (9).

A model called the Actions - Means - Purpose model (The A-M-P model) describes how human trafficking occurs (9). Actions that may be taken by the perpetrator/trafficker include recruiting, harboring, obtaining, patronizing, soliciting, or transporting, while the means include force, coercion or fraud, and the purpose of trafficking is usually sex trafficking (commercial sex) or labor/service trafficking (8). Concerning the dimensional aspects of the means in the A-M-P model, the force could be through physical assault, confinement, or drugging the victim, fraud could be through false promises, fake travel documents, or false advertising, while coercion could be either physical (e.g., threatening to hurt someone, threatening by a weapon) or psychological (e.g., threats against the victim or the family, threats to be sent to jail) (9). Also, human trafficking as a concept differs from human smuggling as they are considered two distinct terms and crimes. While human smuggling requires the movement of victims across borders, human trafficking can occur without leaving the country (10). Although both concepts differ, smuggled humans have a high vulnerability for being victims of human trafficking too (10).

Trafficking of humans results in significant negative impacts on both mental and physical health due to the accompanying physical and psychological violence, in addition to poor living conditions (11). Various psychological responses have been reported by victims including posttraumatic stress symptomology, depression, anxiety, aggression, irritability, poor sleep, insomnia, avoidance and isolation, phobias and panic attacks, substance abuse, hopelessness, distrust and fear of strangers, loss of interest in things, inability to plan for the future, fear of being permanently damaged, and fear of rejection (12).

In addition to the mental health impacts, human trafficking imposes detrimental consequences on victims’ physical well-being too. Victims of human trafficking do not have sufficient access to quality medical care due to various reasons, with financial constraints and the victim’s illegal status being the most common barriers (11). Moreover, victims are more prone to food deprivation and poor nutrition, memory loss, chronic pain, infectious diseases including viral, parasitic, and bacterial diseases (e.g., hepatitis, tuberculosis, sexually transmitted infections, contagious skin infestations), and occupational injuries (12).

Human trafficking is considered a hidden global problem with unsubstantiated numbers despite the presence of some global estimates and reports (13). In 2017, the International Labour Organization (ILO) estimated the presence of approximately 40.3 million victims of human trafficking, of whom 29.4 million were identified as victims of forced labor trafficking (6). Given the sensitivity, and negative consequences of human trafficking on the global system and victims, and considering the scarce research in this area, our current study aimed at analyzing part of the largest anonymized dataset on victims of human trafficking in the period 2010-2020. More specifically, we aimed at describing the following: (i) Sociodemographic profiles of anonymized victims, (ii) Means of control, and (iii) Purpose of trafficking. More details about the dataset and the analysed variables are described in the methods section of this paper.

## 2. METHODS

### 2.1 Study Design

This is a retrospective secondary analysis of a publicly available dataset on global victims of human trafficking.

### 2.2 An Overview of the Dataset and Data Contributors

The dataset used in our analysis is based on the Counter-Trafficking Data Collaborative (CTDC) which is a collaborative hub on human trafficking that provides data through collaboration between various counter-trafficking agencies including the IOM, Polaris, Recollectiv (formerly: the Liberty Shared), The Portuguese Observatory on Trafficking in Human Beings (OTSH), and the A21 (14). Additionally, Microsoft^®^ and Tech Against Trafficking are the technology contributors. The CTDC initiative’s donors are the U.S. Department of State Bureau of Population, Refugees, and Migration (PRM); the U.S. Department of Labour (DOL) through the International Labour Organization (ILO)’s Research to Action (RTA) project; the Global Fund to End Modern Slavery (GFEMS) under a cooperative agreement with the U.S. Department of State (DOS); the IOM Development Fund; and the Ministry of Foreign Affairs of the Netherlands (BZ) (14).

The created dataset by the CTDC initiative is called *the global victim of trafficking dataset*. It is considered the largest global dataset on victims of human trafficking. The required data are collected and updated by the contributing agencies and through the case management system (15). Due to the sensitivity of human trafficking data, individual-level data were protected at two stages, the first stage is through the elimination of all personal identifiers from the dataset, and the second stage which corresponds to a more advanced level of protection is through a mathematical approach called k-anonymization (15). The global K-Anonymized dataset that has been used in our analysis comprises data on more than 156,000 victims of human trafficking from around 189 countries from 2002 to 2021. This dataset used in our analysis is dated 15 July 2021, and it is publicly available to researchers and policymakers through the CTDC website (https://www.ctdatacollaborative.org/dataset/global-k-anonymized-data-and-resources).

### 2.3 Study Population in the Analysed Period

The authors of the present paper have decided to analyze the data for the period 2010 to 2020 using the k-Anonymized global victim of trafficking dataset. In the mentioned period, 87003 victims of human trafficking were identified.

### 2.4 Description of the Analysed Variables

#### 2.4.1 Sociodemographic Information

This part comprised variables that are related to the year of registration (reflects the year in which the victim was identified and assisted by the counter-trafficking agency), data source (reflects the data collection method, whether through case management system or hotline service), gender (female, male, or transgender), age category in years (0-8, 9-17, 18-20, 21-23, 24-26, 27-29, 30-38, 39-47, 48+), majority status at registration (reflects whether the victim was above 18 years or under 18 years of age at the time of registration and assistance by the concerned counter-trafficking agency; thus, categorized as adult or minor), majority Status at exploitation/trafficking (reflects whether the victim was above 18 years or under 18 years of age at the time of trafficking or exploitation; thus, categorized as adult or minor), the victim’s nationality or the country of origin, and country of trafficking/exploitation (the country where a victim is first identified and assisted). Concerning gender classifications, the IOM started to include gender (not sex at birth) including the transgender/non-Conforming category in the survey questionnaire starting in 2017.

#### 2.4.2 Means of Control

In this part, the means of controlling human trafficking victims are collected, and it includes control by debt bondage, control by taking the remunerations/earnings of the victims, control through restrictions of victim’s capability to access the needed daily living funds, control through threats, control through psychological abuse, control through physical abuse, control through sexual abuse, control through false promises, control through psychoactive substances, control through restriction of victim’s freedom of movement, control through restriction of access to medical care, control through excessive working hours, control through restricting the victims’ access to their children, control though threats of law enforcement and imprisonment, control through withholding necessities such as access to basic daily needs, and control through withholding or destroying official documents such as passports, work permits, or other legal or governmental documents. It is important to note that traffickers may use a combination of these means of control to exploit and enslave their victims.

#### 2.4.3 Purpose of Trafficking

This part reflects the purpose of trafficking of the victims, and it includes, forced labor, sexual exploitation and services, combined sex and labor services, forced marriage (a marriage that was imposed on the individual under the threat of a penalty), forced military service (military service that was imposed on the individual under the threat of a penalty), organ removal, as well as slavery and similar practices. Regarding sex services, this included prostitution, pornography, remote interactive services, and private sexual services.

### 2.5 Data Analyses

Data from the k-anonymized global victim of trafficking dataset were extracted and exported to Statistical Package for Social Sciences, SPSS® version 27.0 for Windows (IBM Corp. Version 27.0. Armonk, NY) for quality check and analysis. Descriptive statistics were performed for categorical and numerical data. The age was presented in categories that were described in section 2.4.1. Other categorical variables were presented as frequency counts and percentages. The dataset has many missing values in various parts which represent data that were not collected/missed/could not be retrieved. Missing values were reported where appropriate.

### 2.6 Ethical Considerations

The utilized dataset was already anonymized by the CTDC’s technological support agencies using a two-step anonymization. Additionally, there was no required ethical approval for the utilization of the dataset as it is freely and publicly available to researchers and policymakers through the CTDC initiative website (https://www.ctdatacollaborative.org/dataset/global-k-anonymized-data-and-resources).

## 3. RESULTS

### 3.1 Sociodemographic Profile of Victims

A total of 87003 victims of human trafficking were identified in the period from 2010 to 2020 within the utilized dataset. The most age category encountered among victims was 9-17 years with around 10326 victims (11.9%), followed by 30-38 years with 8562 victims (9.8%). 43656 victims (50.2%) were identified as adults (aged 18 years and above) at the time of registration for assistance by anti-trafficking agencies, whereas 6675 victims (7.7%) were identified as adults at the time of trafficking or exploitation. Females comprised 70% of the sample with 60938 victims. Approximately, 60.5% of the trafficking data were obtained through hotline services. **Table 1** presents more details about the sociodemographic characteristics of the victims.

**Table 1.**
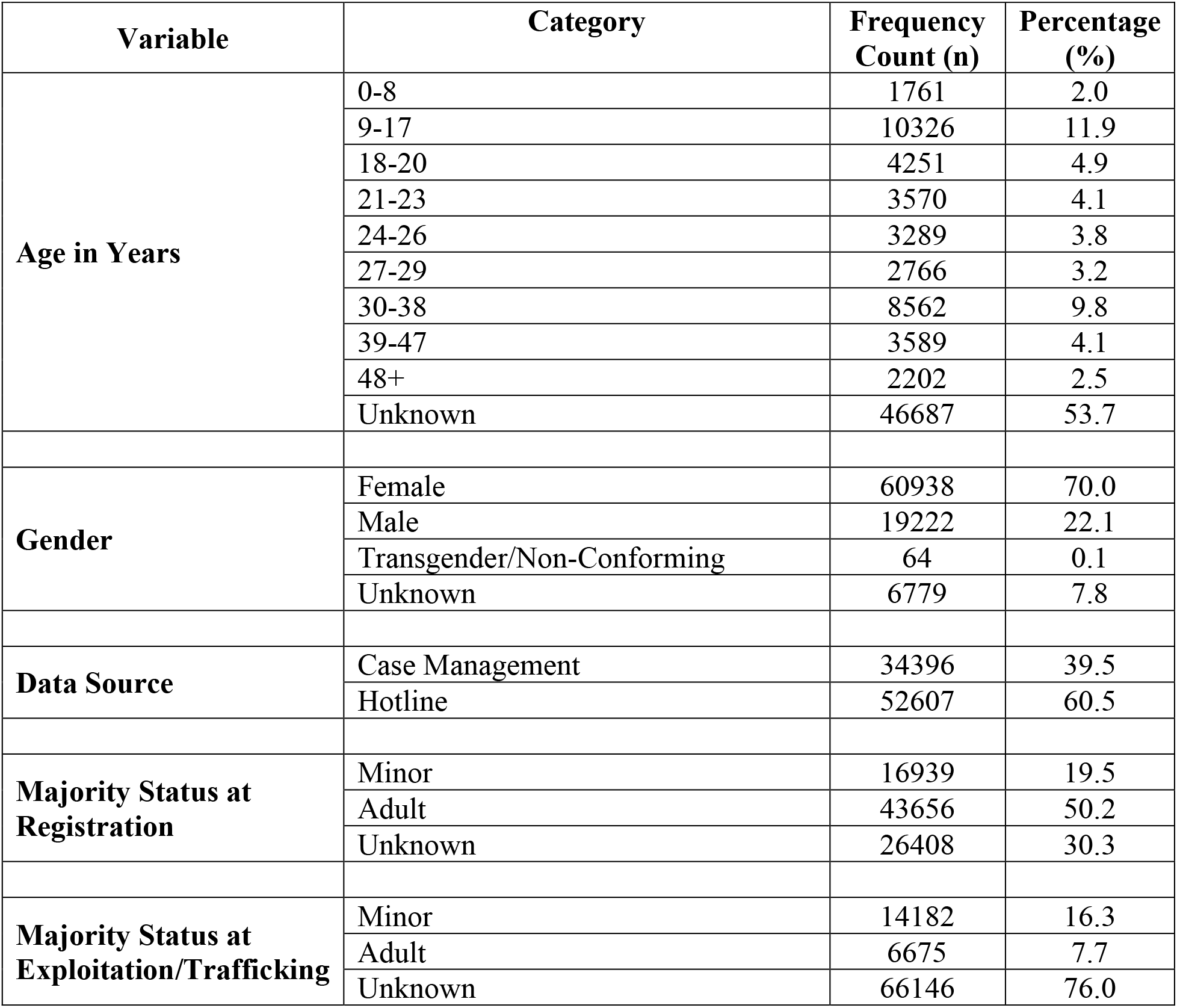
Sociodemographic Profile of Victims in the Period 2010-2020 (n=87003)

Concerning nationality or country of origin of victims, the top 10 countries -in descending order-were the Philippines (n=11454), Ukraine (n=7478), U.S (n=6366), Moldova (n=4984), Mexico (n=2382), Cambodia (n=2130), Indonesia (n=1513), Myanmar (n=1371), Nigeria (n=1040), and Belarus (n=884). 43101 victims (49.5%) had an unknown country of origin (missing value). On the other hand, the top 10 countries of exploitation/trafficking -in descending order-were the U.S. (n=51611), Russia (n=4570), the Philippines (n=1988), Indonesia (n=1784), Cambodia (n=1029), Mali (n=550), Thailand (n=505), Malaysia (n=501), Poland (n=485), and Saudi Arabia (n=355). Missing values related to the country of exploitation were (19072; 21.9%). The year 2019 witnessed the greatest number of victims registered for assistance with around 21312 victims (24.5%), followed by 2016 (n= 18836; 21.6%), and 2018 (n= 18265; 21.0%). See **Figure 1**.

**Figure 1.**
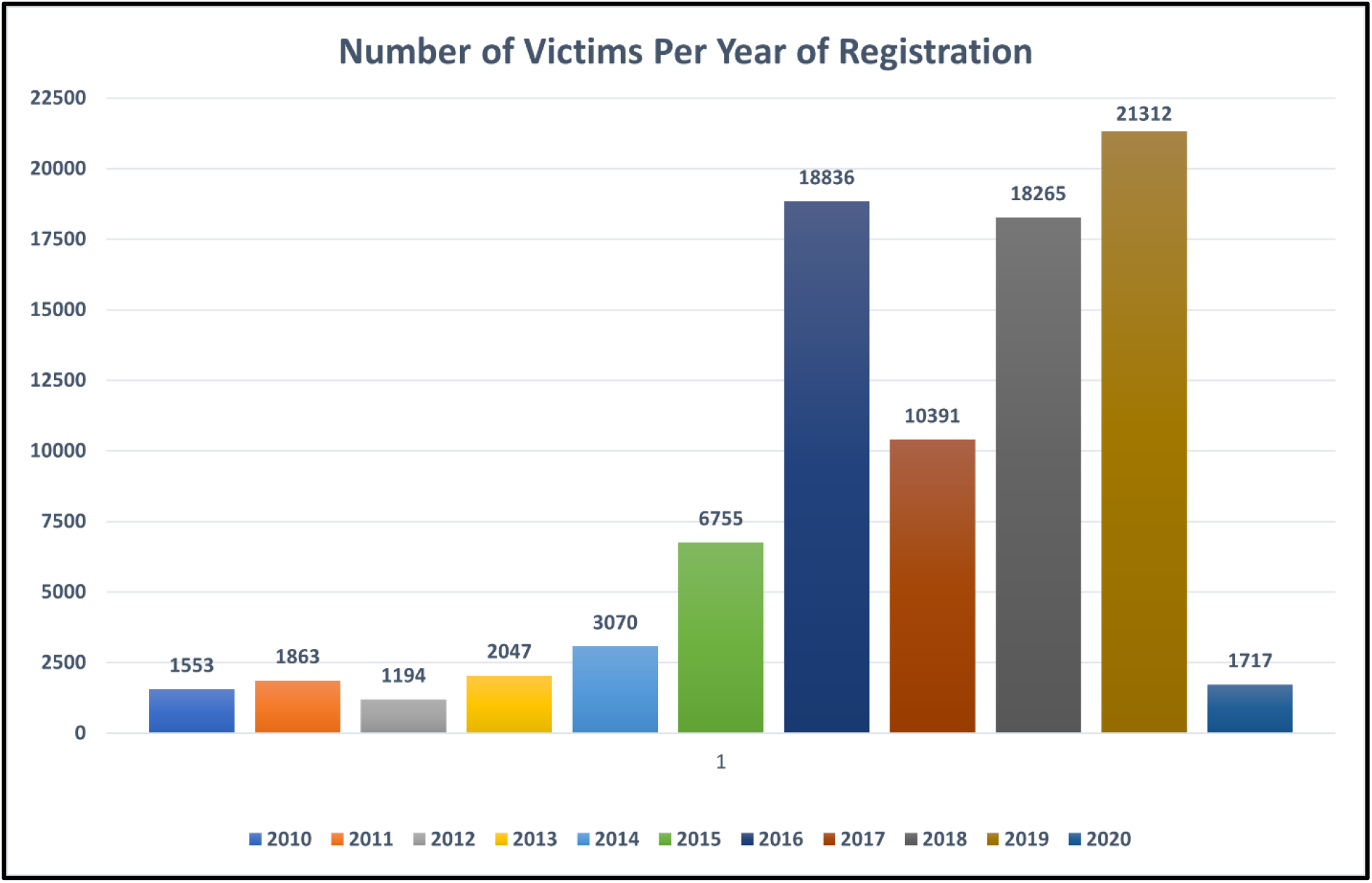
The number of victims per year of registration for assistance (n=87003)

### 3.2 Means of Control

The most reported means of control were threats (n=12343), psychological abuse (n=11980), restriction of the victim’s freedom of movement (n=11059), taking the victim’s remunerations/earnings (n=8588), physical abuse (n=8187), whereas the lowest reported means of control was restricting victims’ access to their children (n=360). **Table 2** shows more details about the reported means of control.

**Table 2.** Means of Control of Victims*

| Mean of Control | Frequency Counts (n) | Percentage (%) |
| --- | --- | --- |
| Threats | 12343 | 14.2 |
| Psychological abuse | 11980 | 13.8 |
| Restriction of victim's freedom of movement | 11059 | 12.7 |
| Taking the victim's remunerations/earnings | 8588 | 9.9 |
| Physical abuse | 8187 | 9.4 |
| Psychoactive substances | 5969 | 6.9 |
| Excessive working hours | 5959 | 6.8 |
| False promises | 5806 | 6.7 |
| Withholding or destroying official documents or governmental documents | 4925 | 5.7 |
| Withholding necessities such as access to basic daily needs | 4813 | 5.5 |
| Threats of law enforcement and imprisonment | 4618 | 5.3 |
| Sexual abuse | 3956 | 4.5 |
| Debt Bondage | 3877 | 4.4 |
| Restriction of access to medical care | 2121 | 2.4 |
| Restrictions on victim's access to daily living funds | 573 | 0.7 |
| Restricting victims' access to their children | 360 | 0.4 |
| Others or Not Specified | 49636 | 57.1 |

\*Multiple means of control can be experienced by the victim

### 3.3 Purpose of Trafficking

Around 42685 victims (49.1%) reported sexual exploitation as the purpose of their trafficking, followed by forced labor with 18176 victims (20.9%). See **Figure 2** for more details. Concerning types of sex work for those who experienced sexual exploitation as a purpose of trafficking were prostitution (n=13287; 15.3%), followed by pornography (n=1866; 2.1%), private sexual services (n=487; 0.6%), and remote interactive sex services (n=118; 0.1%).

**Figure 2.**
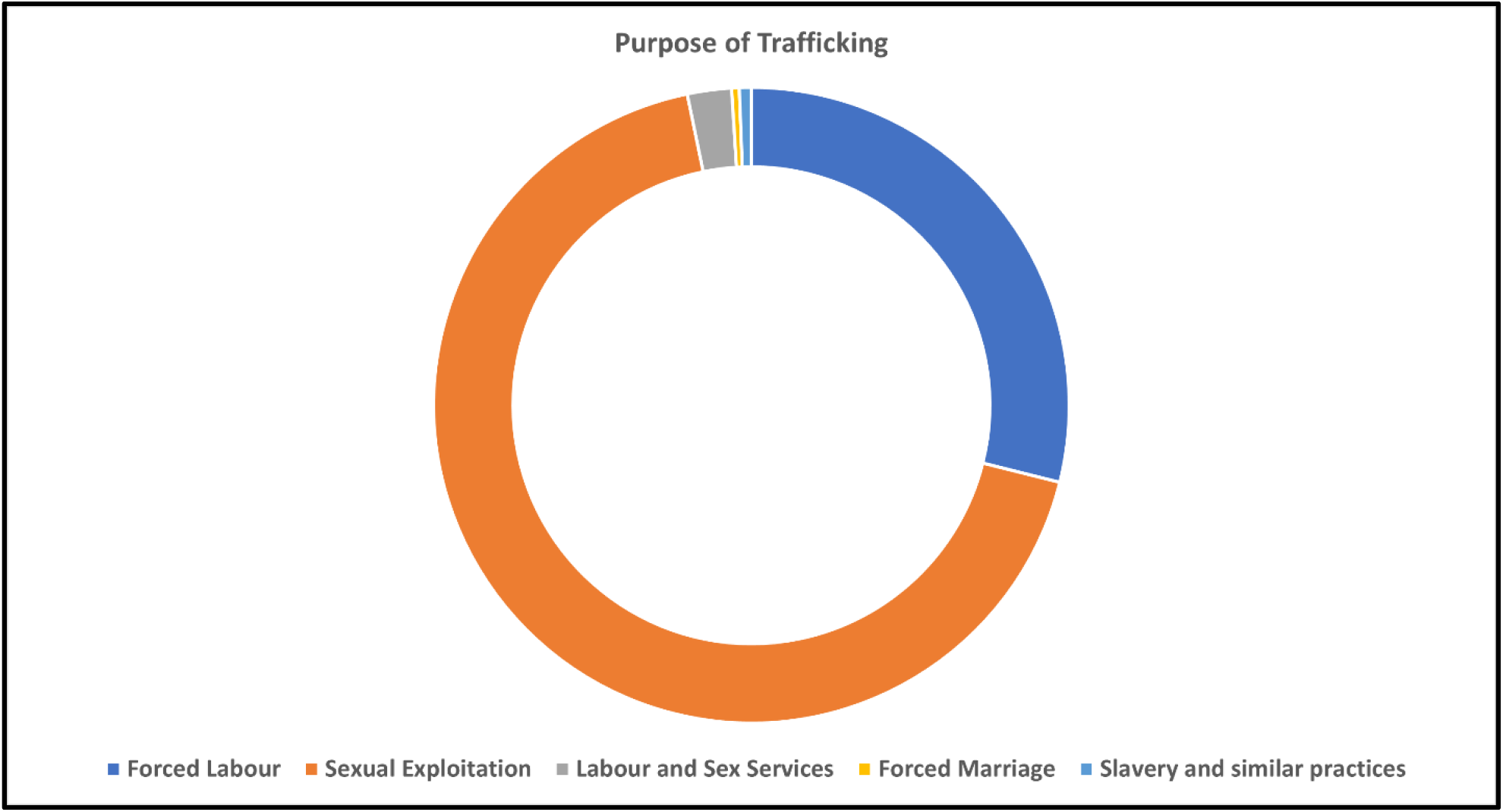
Purpose of Trafficking

## 4. DISCUSSION

To the best of the authors’ knowledge, this is the first scholarly attempt to analyze the largest publicly available data pool on human trafficking in the period 2010-2020, utilizing the K-Anonymized CTDC global victim of human trafficking dataset. The current analysis aimed to describe the anonymized sociodemographic profile of victims, means of control, and the purpose of trafficking. Our paper provides a brief and initial insight into a very sensitive and complicated topic that is of global concern. As mentioned earlier, global migration is affected and shaped by many geopolitical and socioeconomic factors (3,16). These factors may have a negative impact on regular migration (i.e., migration through a legal process) leading to an increased level of irregular migration (i.e., migration through illegal ways). Human trafficking or trafficking in person has been a global concern as it attracted public attention for the past 20 years due to its detrimental and harmful impact on both the mental and physical well-being of victims, its violation of the victims’ dignity, freedom and human rights, as well as its impact on global security (17,18). There are many hazards associated with human trafficking that can be linked to exploitation-related risks (e.g., abuse, occupational hazards, unsafe living conditions, and poor nutrition), or global health (vulnerability to infectious and non-communicable diseases, occupational injuries, financial insecurity, mental health burden, and hazardous labor cycles) (6).

To combat human trafficking, governments and organizations around the world have implemented a range of measures and strategies to prevent trafficking, protect victims, and prosecute traffickers (19). These efforts may include (i) legal frameworks through laws and regulations to criminalize human trafficking and provide protections for victims, (ii) prevention efforts through awareness campaigns, education programs, and efforts to disrupt the demand for exploitation, (iii) protection and support for victims through providing safe and secure housing, medical and mental health care, and legal assistance to victims of trafficking, (iii) prosecution of traffickers through law enforcement and efforts to strengthen the criminal justice system to better respond to trafficking, and (iv) international cooperation through working together and sharing information and resources (20,21). This may involve collaboration across borders and between different sectors, such as the law sector, social services, and non-governmental organizations (NGOs). Additionally, global anti-trafficking efforts may include intelligence and information sharing through gathering and sharing information about human trafficking cases, trends, and networks, as well as using this information to inform law enforcement and victim services efforts (20).

Some examples of global anti-trafficking initiatives and laws are, a collaboration between the IOM, the European Union (EU), the United Nations Office on Drugs and Crime (UNODC) was established to counteract human trafficking through The Global Action against Trafficking in Persons and the Smuggling of Migrants - Asia and the Middle East (GLO.ACT Asia and the Middle East) in the period 2015-2019 (22). Additionally, the UNODC has set a Global Initiative to Fight Human Trafficking (UN.GIFT) which was implemented in 2007 with the main objective of fighting human trafficking through raising public awareness, strengthening preventive measures, supporting and protecting victims, anti-trafficking law enforcement, implementing international commitment, strengthening partnerships, and providing funds for necessary services and initiatives (23). Moreover, various anti-trafficking laws were established in various regions such as the 2006 Ouagadougou Action Plan to Combat Trafficking in Human Beings in the African Union, the Plan of Action Against Trafficking in Persons especially Women and Children - 2015, the Convention Against Trafficking in Persons, Especially Women and Children - 2015, and the Declaration Against Trafficking in Persons, Particularly Women and Children - 2004 which were established by the association of southeast Asian nations (24). The EU has been also active in implementing various measures to fight human trafficking through the 2012-2016 EU Strategy towards the Eradication of Trafficking in Human Beings, and 2011 Directive 2011/36/EU on Combating and Preventing Trafficking in Human Beings and Protecting its Victims. Furthermore, Arab states have joined the global anti-trafficking efforts through the 2012 Comprehensive Arab Strategy for Combating Trafficking in Human Beings, the 2010 Arab Initiative to Combat Trafficking in Persons, and the 2008 Arab Framework Act on Combating Trafficking in Persons (24). The Americas as well have paid tremendous attention to the issue of human trafficking through the 2018 Hemispheric Efforts against Trafficking in Persons “Declaration of Mexico”, the 2014 Inter-American Declaration against Trafficking in Persons, and the 2010-2014 Working Plan to Combat Trafficking in Persons in the Western Hemisphere (24).

Looking at how various countries and nations are standing against human trafficking through various laws and initiatives, it is important to stress the point that anti-trafficking efforts are challenging due to the many complexities surrounding this issue. The UN has classified human trafficking as the third most profitable crime globally (25). Also, human trafficking does not only have economic perspectives but also a public health one. The economic, social, and health impacts of human trafficking are still not thoroughly looked at and researched. According to recent statistics, human trafficking was estimated to be a $150 billion industry (26). As mentioned previously, the ILO revealed that around 40 million people were victimized through trafficking in persons (6). Additionally, the same ILO statistics reported that most victims are females, which is also similar to what was found in our analysis. Also, the findings of our present study are consistent with the 2016 UNODC report which revealed that the majority of trafficked victims were women, girls, and younger children and accounted for 71% (27). Human trafficking is affecting various populations regardless of age, gender, and nationality. Children/minors are also in danger of this crime as they can be trafficked for forced labor, forced military service, and sex trafficking (28). In addition, certain populations have high vulnerability to being victimized through human trafficking such as minors, victims of child sexual abuse, foreign nationals, persons with extreme poverty, and lesbian, gay, bisexual, transgender, queer, and others (LGBTQ+) individuals (28).

To successfully combat human trafficking, anti-trafficking efforts should be brought together in solidarity through inter-governmental and intersectoral partnerships involving governmental organizations, NGOs, the private sector, and communities (29). Tackling a global threat like human trafficking does not have a smooth pathway. Therefore, a multi-faceted approach is required such as the 4Ps paradigm which includes: Protection, Prosecution, Prevention, and Partnerships is considered an essential approach to combat human trafficking (30).

Our study has limitations that should be taken into consideration while interpreting the findings. The CTDC K-Anonymized dataset has many missing values which may change some statistical figures if these missing values were collected. Additionally, there is a possibility of recall bias that impacted the collected data and may have led to the missing values as victims of human trafficking might suffer from psychological traumas that negatively affect their ability to recall events. Another limitation to this study could be that the numbers of those exposed to human trafficking are underestimated and underreported so we cannot assume that the results are generalizable to the population of global victims or international migrants. Nevertheless, our findings provide an initial insight into some statistics related to human trafficking utilizing the CTDC collaborative hub, and we encourage further research in this field with the involvement of a longer period of analysis and utilizing more variables as well.

## 5. CONCLUSION

Human trafficking has detrimental consequences on the physical and psychological health of victims. Various means and methods can be used by traffickers to control the victims to be trafficked for many purposes with sexual exploitation and forced labor being the most common ones. Global anti-trafficking efforts should be brought together in solidarity through utilizing the paradigm of protection of victims, prosecution of traffickers, prevention of trafficking, and inter-sectoral partnerships. Despite being a global concern with various reports that tried to estimate the number of trafficked victims globally, human trafficking still has many unseen aspects that impose a significant challenge and adds to the global burden in combatting this worldwide threat.

## Data Availability

All data produced and analyzed in the current study are available online at:
https://www.ctdatacollaborative.org/dataset/global-k-anonymized-data-and-resources

## DECLARATIONS

### Competing interests

The authors declare that they have no competing interests.

## Acknowledgments

We would like to thank the CTDC initiative and collaborators for providing the anonymized dataset publicly to researchers and policymakers.

## Author Contributions

Conceptualization: ABA; Methodology ABA; Data Analysis: ABA; Supervision: ABA, AN, LK, MS. Writing – original draft preparation: ABA; Writing – literature review, critical review, and editing: ABA, AN, LK, MA, KA, MHA, RA, RS, RA, MS; Visualization: ABA. All authors have significantly contributed to this manuscript. All authors have read and approved the submission of the manuscript.

## Funding

This research received no specific grant from any funding agency in the public, commercial, or not-for-profit sectors.

## Availability of data and materials

The dataset analyzed in the current study is freely and publicly available through this link https://www.ctdatacollaborative.org/dataset/global-k-anonymized-data-and-resources

## Ethics approval and consent to participate

Not required

## Disclaimer

What is expressed in this article is solely those of the authors and does not necessarily represent those of their affiliated organizations.

## Notes

### Competing Interest Statement

The authors have declared no competing interest.

### Funding Statement

This study did not receive any funding

### Author Declarations

The study used ONLY openly available human data that were originally located at: https://www.ctdatacollaborative.org/dataset/global-k-anonymized-data-and-resources

